# Digital Health and Data Utilisation for Improved Primary Health Services Delivery: Multi-Site Perspectives from Quality Improvement Teams in Council Hospitals in Tanzania

**DOI:** 10.64898/2026.04.10.26350674

**Authors:** Constantine Robert Matimo, Godfrey Kacholi, Henry Abraham Mollel

**Affiliations:** Regional Commissioner’s Office, Rukwa, Tanzania; Department of Health System Management, School of Public Administration and Management, Mzumbe University, Tanzania; Centre of Excellence in Health Monitoring and Evaluation, Mzumbe University, Tanzania; Mzumbe University Mbeya Campus College, Mzumbe University, Tanzania

**Keywords:** Digital health technology, Health data use, Improvement of health service delivery, Council Hospitals, Tanzania

## Abstract

**Background:** Digital health plays an indispensable role in facilitating data analysis and use for enhancing healthcare delivery across health settings. However, there is scant information on the extent to which digital health influences the improvement of primary health services delivery through data use. This study examined the determinants that influence the use of digital health to improve health service delivery in council hospitals in Tanzania.

**Methods:** A cross-sectional design was employed in six regions, involving 12 council hospitals. We used a self-administered questionnaire to collect data from 203 members of hospital quality improvement teams. Descriptive analysis was used to determine the frequency, proportion, and mean of responses, while bootstrapping analysis was conducted to test the statistically significant influence of digital health factors on data use for improving health service delivery.

**Results:** Results show moderate agreement on data compatibility for planning and decision-making, with 40.4% of respondents agreeing it supports ordering commodities, 43.8% for staff allocation, and 38.4% for planning. However, dissatisfaction was higher for user-friendliness (47.8%), reliability (up to 65.5%), and usefulness (up to 63.5%). Overall, 50.2% (M=2.74±0.87) disagreed that digital systems effectively support data use. Structural model analysis confirmed significant positive influence of usefulness (β=0.199, p<0.001) and access to quality data (β=0.729, p<0.001) on data use, which strongly impacted service delivery (β=0.593, p<0.001), despite some factors showing no direct influence.

**Conclusion:** The study finds that current digital health initiatives only modestly improve the user-friendliness, reliability, and usefulness of data systems, partly due to fragmented, non–interoperable platforms that burden data management. However, compatibility, usability, reliability, and usefulness of digital tools significantly enhance access to quality data and data-driven decisions. The study recommends strengthening and integrating existing systems and providing continuous digital health training to institutionalize data-informed decision-making.

## Background

Health data utilization is a crucial component of the healthcare system, enabling the development of evidence-based plans, informed resource allocation, and informed decision-making. The health system monitors a range of health services through data collection and use for decision-making in improving health service delivery [1]. Health data use is a process that involves accessibility to quality data and data application for planning and decision-making [2]. Data visualization and information sharing with stakeholders through dashboards, data dissemination through notice boards, and data review and interpretation are examples of access to quality data [3]. Further, ordering essential health commodities using existing data systems, allocating hospital staff based on patient load, and preparing hospital plans and budget allocation based on existing data systems are examples of data applications for planning and decision-making [4].

However, there exists a global trend of inadequate utilization of health data by policymakers and decision-makers, including health managers, leading to compromised efficiency, accessibility, and quality in health service delivery [5]. Despite significant investments in strengthening data systems and data generation in Sub-Saharan countries, including Tanzania, challenges persist regarding access to quality data and data use for planning and decision-making [6]. These challenges have highlighted the gaps between data availability and its utilization for planning and decision-making processes [1,4,6].

Attaining adequate health data use requires the influence of several associated factors, such as organizational and digital health factors within healthcare settings [6,7] . Previous studies in low- and middle-income countries have highlighted the significant negative impact of various digital health factors [8,9]. These factors have consequently impacted the utilization of health data in healthcare settings [10]. In Tanzania, efforts have been made through the implementation of eHealth strategies and the connection of DHIS-2 to all Local Government Authorities (LGAs) in primary health facilities for data collection, analysis, and use [11,12]. However, there is plenty of data in digital health systems and paper-based systems that are underutilized for planning and decision-making to improve health service delivery in healthcare settings [13,14].

Digital health encompasses the use of information and communication technologies (ICTs) to support and improve health and healthcare [10]. It involves a broad range of technologies, including mobile health (mHealth), telehealth and telemedicine, wearable devices, Health Information Technology (HIT), and personalized medicine [15]. Digital health technology encompasses the specialized knowledge and technological development, design, management, and improvement of data analysis, reporting, and the use of information necessary for monitoring and evaluating targeted health interventions. Additionally, previous studies demonstrated that the influence of modern information technology facilitates an environment conducive to adequate health data use and improved health service delivery in terms of compatibility, usability, reliability, and usefulness of IT [10,16].

### Theoretical Perspective

The Unified Theory of Acceptance and Use of Technology (UTAUT) framework was utilized to deepen the analysis of how various digital health factors affect the utilization of data within healthcare systems. The application of the UTAUT aimed to clarify the dynamics influencing data access and usage, particularly about supporting planning and evidence-based decision-making processes essential for enhancing the quality of health service delivery (Figure 1).

**Figure 1:**
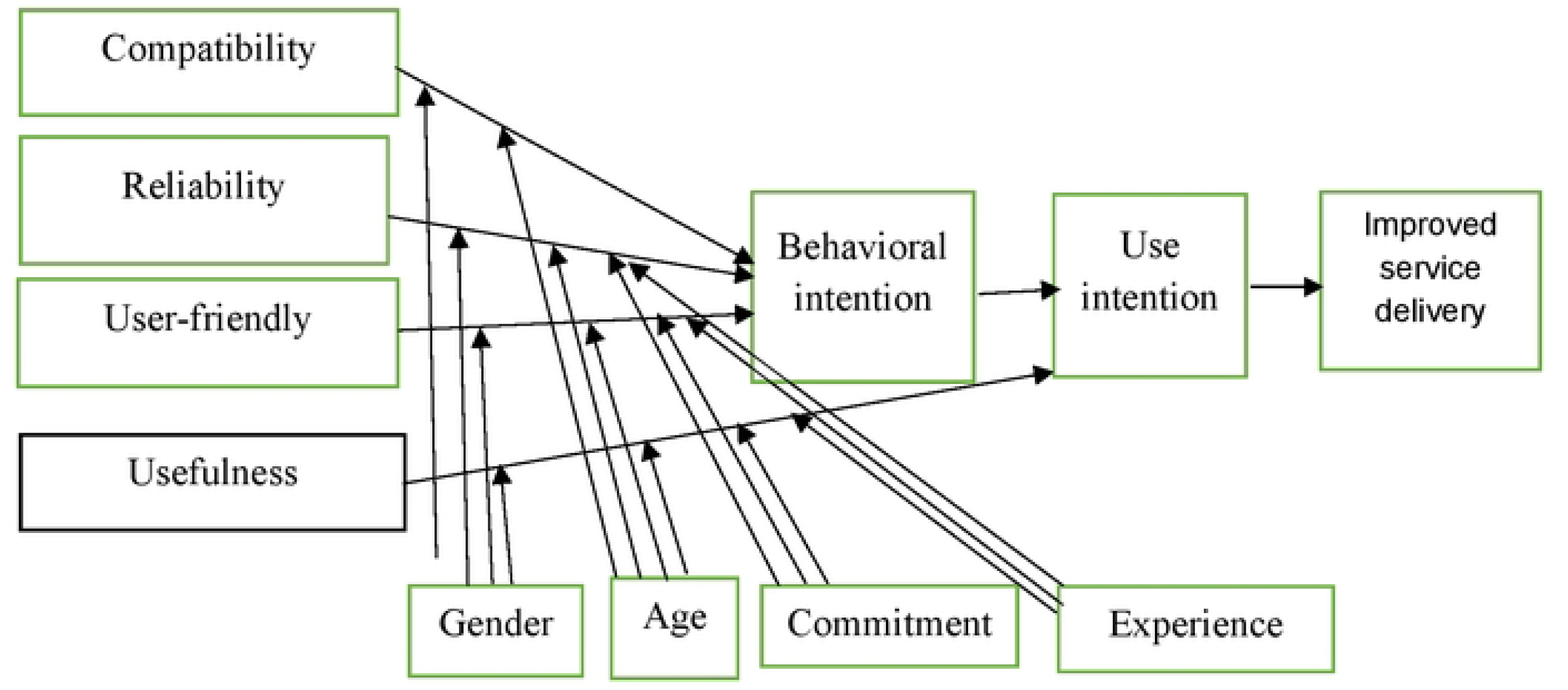
The Modified UTAUT Models on Digital Health Factors Influence Data Use for Improvement of Health Service Delivery (Venkatesh et al., 2016)

The four moderating variables (compatibility, reliability, user-friendliness and usefulness) were assumed to moderate the influence of the four core constructs on behavioural intention and use intention for improved health service delivery. These relations have been modified and summarized from the hypotheses of [16,17] studies as follows: The compatibility of a computerized system captures the extent to which an innovation is consistent with the existing technical and social environment [17]. Roger’s classical innovation diffusion theory provides a linkage between computerized systems and adoption decisions, describing five attributes specific to technological innovation: user-perceived qualities, relative advantage, compatibility, complexity, trialability, and observability. Compatibility has been recognized as a key factor influencing the adoption and use of computerized systems for decision-making[18]. User-friendly computerized systems play a crucial role in facilitating rapid software understanding and influencing the environment for health data effectiveness [16]. End users are more likely to rapidly accept a digital technology that is less complex [19,20]. However, other studies suggest that complex software negatively affects data use, as the complexity of digital health technologies gauges the level of difficulty health workers perceive in understanding, implementing, or using an innovation [8,9,21].

The reliability of computerized systems is essential in enabling an environment for health data use. According to [8], reliability refers to the degree to which users believe that using technologies will result in improving service delivery. In contrast, a system with slow response times, inaccurate and incomplete outputs, or frequent crashes has detrimental effects on user att. If users spend more time on a system and gain nothing from it, they are likely to feel distressed eventually. Conversely, users tend to use information systems intensively if they provide timely, complete, and accurate outputs that meet their needs and facilitate the achievement of their goals [22].

The usefulness of a computerized system refers to the degree to which the user perceives benefits or improvements to the existing technology by adopting an innovation [16]. For instance, [23] conducted a study using a model that incorporated the UTAUT to confirm the degree of acceptance of e-health applications among German healthcare professionals. They found that perceived usefulness (PU) had a positive influence on the intention to use the existing data system in a health facility. Several scholars [9,15,24] have acknowledged the importance of perceived usefulness in influencing high acceptance of IT. Likewise, [8] also revealed that perceived usefulness is a crucial factor influencing the adoption and effectiveness of information systems.

In sub-Saharan countries, including Tanzania, significant investments have been made in data systems and data generation through existing data systems. However, there is scant information on the extent to which digital health factors influence the effective use of health data to improve health service delivery in healthcare settings. This study examined the determinants that influence the use of digital health to improve health service delivery in council hospitals in Tanzania. Consequently, the following hypotheses were tested.

- H1: Digital health factors have a positive and significant influence on access to quality data in council hospitals.
- H2: Access to quality data has a positive and significant influence on data use for planning and decision-making.
- H3: Data use for planning and decision-making has a positive and significant influence on the improvement of health services delivery.
- H4 Access to quality data has a positive and direct significant influence on improvement of health service delivery.
- H5: Digital health factors have a positive and direct significant influence on the improvement of health service delivery.

## Materials and methods

### Study design and setting

A cross-sectional study was conducted across six regions, encompassing twelve council hospitals from 31, March 2023 to 30, March, 2024. The selected hospitals comprised Mbalizi Council Designated Hospital and Chunya District Council Hospital in Mbeya Region; Mjimwema Municipal Council Hospital and Peramiho Council Designated Hospital in Ruvuma Region; Hai and Same District Council Hospitals in Kilimanjaro Region; Magu and Nyamagana District Council Hospitals in Mwanza Region; as well as Bunda and Tarime Town Council Hospitals in Mara Region. Hospital selection was purposive and guided by the national star rating system, incorporating facilities with performance ratings both below and above three stars.[25].

### Sampling size and sampling procedures

A total of 218 members of hospital Quality Improvement Teams (QIT) were selected for the study using the Yamane formula for sample size determination. The calculation was conducted at a 95% confidence level, considering a population proportion (P) of 0.5 to maximize variability. The formula applied was

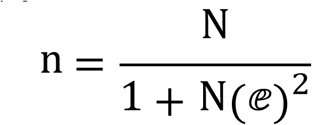

Where *n* represents the sample size, *N* is the population size (in this case, the total number of QIT members across the selected council hospitals), and *e* is the level of precision or margin of error set at 0.05.

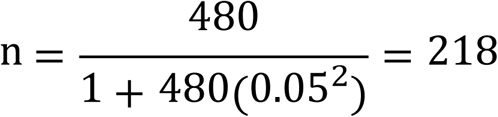

The study employed a multi-stage sampling procedure to select regions, council hospitals, and respondents. In the first stage, stratified sampling was used to select 26 regions: 10 high-performing and 16 low-performing, based on performance criteria. In the second stage, six regions—Songwe, Mbeya, Ruvuma, Kilimanjaro, Mwanza, and Mara—were chosen through simple random sampling, with three selected from each performance category. The third stage involved stratified sampling to select 40 council hospitals from the chosen regions, including 22 high-performing hospitals (rated 3 stars or above) and 18 low-performing hospitals (rated below 3 stars). In the fourth stage, 12 council hospitals—2 from each region—were selected by simple random sampling, ensuring representation of both high- and low-performing facilities. Finally, at the fifth stage, a total of 218 QI team members were selected by simple random sampling, with 109 from high-performing council hospitals and 109 from low-performing hospitals.

### Data collection tools

Data were collected using structured self-administered questionnaires from 218 respondents sampled from selected council hospitals. The questionnaire was designed to capture both socio-demographic characteristics and key variables related to the influence of digital health factors on data use for planning, decision making, and health service delivery improvement. The socio-demographic section gathered information on respondents’ age, sex, marital status, educational qualifications, and professional roles. The core section of the questionnaire was designed to collect respondents’ perceptions on digital health system characteristics, including compatibility, user-friendliness, reliability, and usefulness of existing data systems for various health management functions—such as ordering essential commodities, staff allocation, and planning and budget allocation. Responses were measured using a Likert scale. To complement the perception data, information on the extent and pattern of digital health factors’ influence on data use for planning and decision-making was also collected. Validity and reliability of the constructs were ensured through assessments of indicator loading, internal consistency reliability (Cronbach’s alpha, composite reliability), and convergent/discriminant validity (Average Variance Extracted).

### Data management and analysis

The collected data were reviewed daily for completeness and consistency. The information was then cleaned, edited, and aggregated before being entered into SPSS version 26. Descriptive statistics—including frequencies, proportions, and means—were calculated to summarize responses. SmartPLS 3 was utilized with bootstrapping techniques to test associations between key factors and the use of health data for improving health service delivery. Advanced statistical methods, notably Partial Least Squares Structural Equation Modelling (PLS-SEM), were also applied facilitate the analysis of the relationships among digital health factors, data utilization, access to quality data, and improvements in health service delivery.

### Ethical Approval

The study received ethical approval from the National Institute for Medical Research, Tanzania (Reference: NIMR/HQ/R.8a/Vol.IX/4251), and the Postgraduate Technical Committee of Mzumbe University (Reference: MU/PhD/SOPAM/MZC/040/T.2020). Additional permissions were secured from the respective local government authorities prior to data collection. Informed written consent was obtained from all participants following a comprehensive explanation of the study’s objectives, potential benefits, and associated risks. Participation was entirely voluntary, with all participants retaining the right to withdraw from the study at any point without obligation to provide a reason.

## Results

### Socio-demographic characteristics of respondents

A total of 203/218 (93.1%) members of quality improvement teams were interviewed in this study. The majority (50.7%) of participants were aged 31–40 years. Females comprised a slightly higher proportion of participants (56.2%) compared to males (43.8%). Most participants were married (83.3%). Educational attainment was predominantly at the certificate or diploma level (77.3%), with only 20.7% holding bachelor’s degrees and just 2.0% having Masters or PhD qualifications. The vast majority of surveyed staff were clinical (82.3%), while non-clinical personnel accounted for only 17.7% (Table 1).

**Table 1:** Socio-demographic characteristics of respondents (N=203)

| Variable | Response | Frequency (N) | Percentage (%) |
| --- | --- | --- | --- |
| Age | 21-30 years | 28 | 13.8 |
|  | 31-40 years | 103 | 50.7 |
|  | 41-50 years | 54 | 26.6 |
|  | 51-60 years | 16 | 7.9 |
|  | > 60 years | 2 | 1.0 |
| Sex | Female | 114 | 56.2 |
|  | Male | 89 | 43.8 |
| Marital status | Married | 169 | 83.3 |
|  | Separated | 1 | 0.5 |
|  | Single | 30 | 14.8 |
|  | Widow | 3 | 1.5 |
| Educational qualifications | Certificate/Diploma | 157 | 77.3 |
|  | Bachelor Degree | 42 | 20.7 |
|  | Masters/PhD Degree | 4 | 2.0 |
| Professionalism | Clinical | 167 | 82.3% |
|  | Non-clinical | 36 | 17.7% |

### Digital health-related factors influencing data use

Compatibility was most favourable for ordering essential health commodities (40.4% agreed, mean 3.13±0.84) and for hospital planning and budgeting (38.4% agreed, mean 3.11±0.78). Compatibility was lowest for staff allocation (only 43.8% agreed, mean 2.67±0.82). User-friendliness generally received less positive ratings; between 21.2% and 27.1% agreed that tools were user-friendly for various processes, with mean values ranging from 2.73 to 2.82. Reliability assessments were mostly negative: over half disagreed about the reliability of tools, especially for hospital staff allocation (65.5% disagreed, mean 2.38±0.70). Usefulness was rated modestly across most tasks, with agreement rates mostly below 30%, and mean scores between 2.50 and 2.81. The overall technological factors for data use in planning, decision-making, and management (DUPDM) yielded a low mean (2.74±0.87), with half of the respondents disagreeing that the technology supports these functions well (Table 2).

**Table 2:** Digital health-related factors influencing data use (N=203)

| **Aspects Assessed** | **Disagree**<br>**n (%)** | **Neither agree**<br>**nor disagree**<br>**n (%)** | **Agree**<br>**n (%)** | **Mean**<br>**±SD** |
| --- | --- | --- | --- | --- |
| Compatibility in ordering essential health commodities | 56(27.6) | 64(31.5) | 82(40.4) | 3.13±0.84 |
| Compatibility in allocating hospital staff based on patient load | 69(34.0) | 45(22.2) | 89(43.8) | 2.67±0.82 |
| Compatibility in preparing hospital plans and budget allocation | 51(25.1) | 74(36.5) | 78(38.4) | 3.11±0.78 |
| User-friendliness for ordering essential health commodities | 92(45.3) | 56(27.6) | 55(27.1) | 2.82±0.83 |
| User-friendliness for allocating hospital staff | 96(47.3) | 65(32.0) | 42(20.7) | 2.73±0.78 |
| User-friendliness for preparing hospital plan and budget allocation | 97(47.8) | 63(31.0) | 43(21.2) | 2.73±0.79 |
| Reliability for ordering of essential health commodities | 112(52.2) | 36(17.7) | 55(27.1) | 2.72±0.87 |
| Reliability for allocating hospital staff | 133(65.5) | 27(13.3) | 43(21.2) | 2.38±0.70 |
| Reliability for preparing hospital plans and budget allocation | 128(63.1) | 30(14.8) | 45(22.2) | 2.59±0.83 |
| Usefulness for ordering of essential health commodities | 96(47.3) | 50(24.6) | 57(28.1) | 2.81±0.85 |
| Usefulness for allocating hospital staff | 129(63.5) | 32(15.8) | 42 (20.7) | 2.50±0.90 |
| Usefulness for preparing hospital plans and budget | 111(54.7) | 47(23.2) | 44(21.7) | 2.70±1.63 |
| Overall technological factors for DUPDM | 102(50.2) | 49(24.1) | 51(25.1) | 2.74±0.87 |

### Validation of Constructs Measuring Digital Health, Health Data Use, and Service Delivery

The analysis of the measurement model showed that indicator loadings ranged from 0.580 to 0.980, except for PHPBEDS (0.514), which was retained due to its theoretical relevance. Cronbach’s alpha values ranged from 0.732 to 0.945, all surpassing the 0.70 reliability threshold. Average Variance Extracted (AVE) values were between 0.557 and 0.902, exceeding the recommended cutoff of 0.50—indicating good convergent validity. The study employed PLS-SEM, evaluating both the measurement and structural models to ensure the validity and significance of the relationships among digital health factors, health data use, and service delivery improvement (Table 3).

**Table 3:** Validation of Constructs for Digital Health, Data Use, and Service Delivery.

| Construct | Variable | Indicator | Loading factor | Cronbach's Alpha | composite reliability | (AVE) |
| --- | --- | --- | --- | --- | --- | --- |
| Digital Health factors | COEDS | CDVS | 0.859 | 0.856 | 0.912 | 0.776 |
|  |  | CDDN | 0.930 |  |  |  |
|  |  | CDRIM | 0.852 |  |  |  |
|  | USFEDS | USDVS | 0.894 | 0.894 | 0.934 | 0.826 |
|  |  | USDDN | 0.951 |  |  |  |
|  |  | USDRIM | 0.881 |  |  |  |
|  | REDS | RDVS | 0.892 | 0.888 | 0.930 | 0.816 |
|  |  | RDDN | 0.923 |  |  |  |
|  |  | RDRIM | 0.895 |  |  |  |
|  | USEDs | UDVS | 0.928 | 0.945 | 0.965 | 0.902 |
|  |  | UDDN | 0.966 |  |  |  |
|  |  | URIM | 0.954 |  |  |  |
| Health data use | AQD | DVSMB | 0.960 | 0.920 | 0.949 | 0.862 |
|  |  | DDMB | 0.928 |  |  |  |
|  |  | DRIM | 0.897 |  |  |  |
|  | DUPDM | OEHCBEDS | 0.980 | 0.790 | 0.883 | 0.728 |
|  |  | AHSPSL | 0.979 |  |  |  |
|  |  | PHPBEDS | 0.514 |  |  |  |
| Health services delivery | IPHSD | AEHC | 0.882 | 0.732 | 0.831 | 0.557 |
|  |  | AHSBAR | 0.726 |  |  |  |
|  |  | AEPBLA | 0.580 |  |  |  |
|  |  | AMHIT | 0.765 |  |  |  |

**Variables:** COED=Compatibility of existing data systems, USFEDS= User-friendliness of existing data systems, REDS= Reliability of existing data systems, USDDS= Usefulness of existing data systems, AQD=Access to quality data, DUPDM= Data use for planning and decision making, IPHSD=Improvement of primary health service delivery

### Evaluation of Structural Model

As indicated in Table 4, all three endogenous constructs achieve acceptable levels of explanatory power and predictive accuracy. For explanatory power, access to quality data (AQD) has an R^2^of 0.501 and improvement in health service delivery (IPHSD) has an R^2^ of 0.380, both indicating moderate explained variance within the commonly accepted range of 0.25–0.75. Data use for planning and decision-making (DUPDM) shows a higher R^2^ of 0.780, suggesting substantial explanatory power relative to the other constructs. The associated t-values for all R^2^estimates are high, supporting their statistical significance.

**Table 4:** Explanatory Power (R²) and Predictive Accuracy (Q²) of Endogenous Constructs.

| Endogenous construct | Explanatory predictive power |  |  | Path model of predictive accuracy |  |  | Results |
| --- | --- | --- | --- | --- | --- | --- | --- |
| | $R^2$ | t-test ($R^2$) | Normal $R^2$ range | SSO | SSE | $Q^2$ (=1-SSE/SSO) | |
| AQD | 0.501 | 8.612 | 0.25–0.75 | 609 | 351.668 | 0.423 | Accepted |
| DUPDM | 0.780 | 26.347 | 0.25–0.75 | 609 | 273.042 | 0.552 | Accepted |
| IPHSD | 0.380 | 7.261 | 0.25–0.75 | 812 | 646.977 | 0.203 | Accepted |

*AQD = Access to Quality Data; DUPDM = Data Use for Planning and Decision-Making; IPHSD = Improvement of Health Service Delivery.*

For predictive accuracy, all Q² values are greater than zero, indicating that the model has predictive relevance for each construct. AQD (Q² = 0.423) and DUPDM (Q² = 0.552) exhibit moderate predictive accuracy, while IPHSD (Q² = 0.203) indicates lower but still acceptable predictive relevance. Overall, the results confirm that the structural model provides adequate explanatory power and predictive accuracy for the key endogenous constructs.

### Structural Model Results for Health Information Technology, Data Use, and Service Delivery

As indicated in Table 5, the health information technological factor is operationalized through compatibility, user-friendliness, reliability, and usefulness of existing data systems in relation to access to quality data and data use for planning and decision-making. The table reports that hypotheses H1 (β = 0.145, p = 0.002), H2 (β = 0.540, p < 0.001), H3 (β = −0.259, p < 0.001), H4 (β = 0.369, p < 0.001), and H8 (β = 0.199, p < 0.001) are supported. The table also shows that hypotheses H5 (β = −0.075, p = 0.045), H6 (β = 0.110, p = 0.141), and H7 (β = −0.058, p = 0.368) are rejected. Furthermore, Table 5 presents the assessment of the influence of access to quality data and data use for planning and decision-making on the improvement of primary health service delivery. The results indicate that hypotheses H9 (β = 0.729, p < 0.001) and H11 (β = 0.593, p < 0.001) are supported, while hypothesis H10 (β = 0.100, p = 0.397) is rejected.

**Table 5:** Hypotheses testing the relationship among the constructs.

| Hypothesis | Path in the structural model | Path coefficient ($\beta$) | Significance (P Value) | Results |
| --- | --- | --- | --- | --- |
| H1 | COEDS -> AQD | 0.145 | 0.002 | Supported |
| H2 | USFEDS -> AQD | 0.540 | $P < 0.001$ | Supported |
| H3 | REDS -> AQD | -0.259 | $P < 0.001$ | Supported |
| H4 | USEDSD -> AQD | 0.369 | $P < 0.001$ | Supported |
| H5 | COEDS -> DUPDM | -0.075 | $P = 0.045$ | Rejected |
| H6 | USFEDS -> DUPDM | 0.11 | $P = 0.141$ | Rejected |
| H7 | REDS -> DUPDM | -0.058 | $P = 0.368$ | Rejected |
| H8 | USEDSD -> DUPDM | 0.199 | $P < 0.001$ | Supported |
| H9 | AQD -> DUPDM | 0.729 | $P < 0.001$ | Supported |
| H10 | AQD -> IPHSD | 0.1 | $P = 0.397$ | Rejected |
| H11 | DUPDM -> IPHSD | 0.593 | $P < 0.001$ | Supported |

### Structural Model Pathways for Digital Health Factors and Health Service Delivery Improvement

Figure 2 presents the structural model illustrating the statistically significant and non-significant pathways among the study constructs. The model depicts four dimensions of digital health technology—compatibility (COEDS), user-friendliness (USFEDS), reliability (REDS), and usefulness (USEDS) of existing data systems—as antecedents to access to quality data (AQD) and data use for planning and decision-making (DUPDM). The figure demonstrates both direct and indirect pathways through which these health information technology factors influence the improvement of primary health service delivery (IPHSD) in Tanzanian council hospitals.

**Figure 2:**
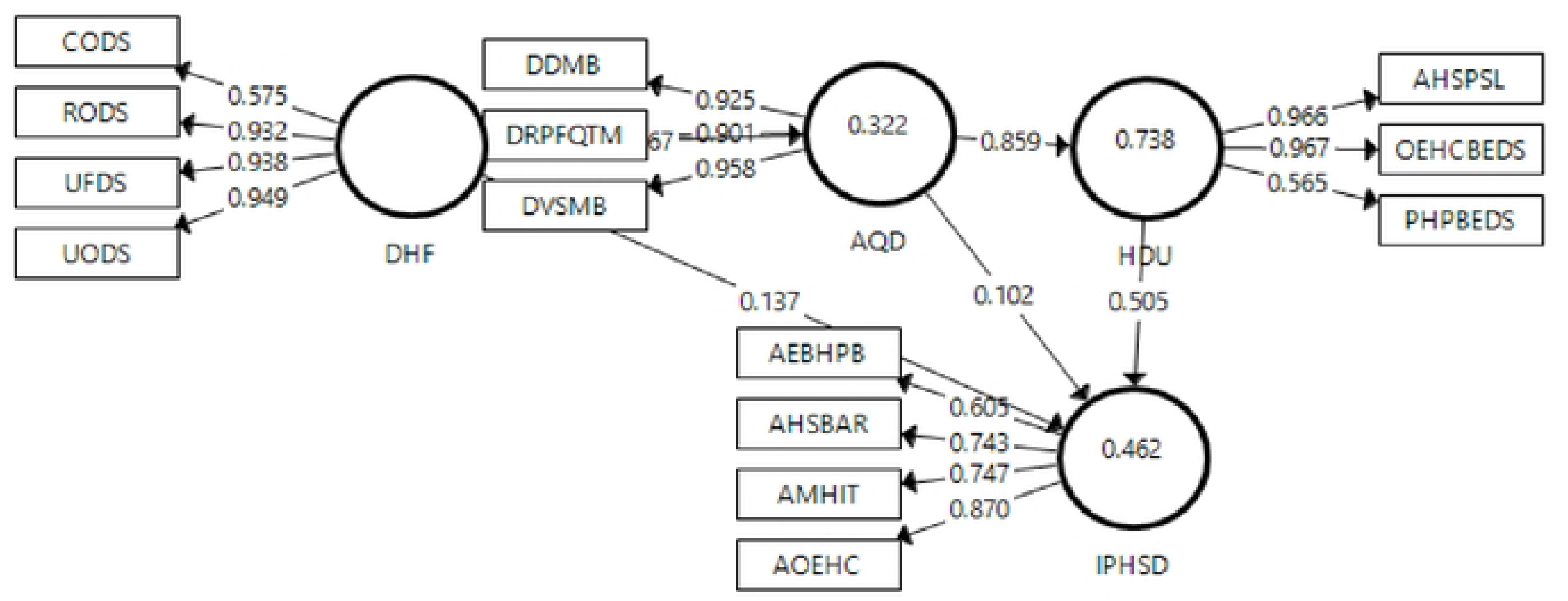
Structural Model of Digital Health Factors, Data Use, and Health Service Delivery Improvement in Tanzanian Council Hospitals

## Discussion

The findings indicate that 44.8% of respondents (mean = 2.83 ± 0.95) and 50.2% of respondents (mean = 2.74 ± 0.87) disagreed that existing digital health systems adequately support access to quality data and data use for planning and decision-making with respect to user-friendliness, reliability, and usefulness. This pattern reflects a generally low perceived usability of digital health systems in council hospitals in Tanzania. In contrast, regarding compatibility, more than half of the respondents (58.0%) agreed that information in existing data systems is aligned with the data used for visualization, dashboard reporting, dissemination, and interpretation. These results are consistent with prior studies, which have shown that perceived compatibility significantly influences the adoption of computerized systems [18] and [17]access to quality data in health facility settings.

The present study reveals that a significant majority of respondents expressed disagreement regarding the user-friendliness of existing data systems for purposes such as data visualization, analysis, dissemination via notice boards, and interpretation. Respondents attributed this dissatisfaction primarily to the limited or inadequate training provided on the use of these systems. Supporting this observation, a prior study indicates that the complexity inherent in digital health technologies can impede health workers’ capacity to comprehend, implement, and effectively utilize these innovations [8] . This finding highlights a critical misalignment between the design of digital health systems and the technical competencies of users, suggesting that the potential of digital health tools is not being fully realized to meet routine information requirements at the facility level.

The study also reveals that a considerable proportion of the respondents expressed low confidence in the reliability of existing data systems for data visualization, sharing through dashboards, dissemination via notice boards, and review through quarterly quality improvement team meetings. These findings are consistent with [26], who reported that the majority of hospital staff found existing digital systems unreliable for data analysis and sharing, attributing this to inaccuracies and incomplete outputs negatively impacted user attitudes. Similarly, [27] found that only 69% of physicians believed electronic health records were not reliable for accessing quality data for medical procedure management in hospitals. These findings underscore the need for council hospitals to improve trust and confidence in their data systems to facilitate informed decision-making based on accurate and reliable data.

Previous studies highlight the significant influence of perceived usefulness on the adoption of digital systems for data visualization, sharing, review, and dissemination to relevant stakeholders in health facility settings. However, a study by [8] indicates that a minority of physicians and nurses found existing modern health information technologies useful for accessing quality data. These findings are consistent with another study indicating that a significant proportion of respondents disagree that existing data systems are useful for data analysis and sharing through dashboards with targeted users, due to inadequate training in data analysis and interpretation within these systems in health facilities.

The study findings also indicate that a considerable proportion of respondents reported low perceived usefulness of existing data systems for accessing quality data through visualization, dashboard-based sharing, dissemination via notice boards, and routine review. In a similar vein, reported that perceived usefulness positively influences healthcare professionals’ acceptance of modern health information technologies for data analysis, interpretation, and targeted information sharing [23]. Taken together, these findings highlight the need for health facilities to strengthen the functional relevance and perceived added value of their data systems, thereby enabling staff to utilize data more effectively in support of evidence-informed decision-making.

In the realm of data utilization for strategic planning and decision-making, the findings reveal that a significant proportion of respondents expresses skepticism regarding the efficacy of existing information technologies, particularly in relation to their compatibility, user-friendliness, reliability, and overall usefulness. Specifically, regarding compatibility, findings indicate that nearly half of the participants affirm that the information in current data systems aligns with the data used for the procurement of essential health commodities in healthcare facilities. This observation is corroborated by the results of the previous study, which underscores the critical importance of accurate data sources and high-quality information in effectively managing and monitoring health commodity supply chains [28]. Such alignment is essential to ensure sufficient quantities of appropriate health commodities are available at service delivery points, thereby effectively addressing patients’ healthcare needs.

The findings presented in this study are consistent with the results demonstrating that a considerable proportion of respondents express incongruity regarding the compatibility of information in existing data systems with the data utilized for the allocation of hospital staff at both department and unit levels. Furthermore, a previous study indicates that multiple registries capturing the same data items—such as patient information, injections, dispensing, antiretroviral therapy (ART), tuberculosis (TB), reproductive and child health (RCH), and prevention of mother-to-child transmission (PMTCT)—may coexist within a single healthcare facility [29]. This redundancy may lead to data incompatibility, subsequently affecting the alignment of information necessary for effective hospital staff allocation. In addition, the study shows that a substantial proportion of respondents did not indicate a clear position on whether existing data systems are compatible with the data used in preparing hospital plans and budget allocations. This neutrality can be attributed to insufficient training provided to hospital staff in the analysis and application of software during the hospital planning processes. These findings are corroborated by previous research that identified capacity constraints, limited computerization, and inadequate internet connectivity in primary health facilities as significant barriers to leveraging existing data systems for planning and budget allocation [11].

The study findings indicate that nearly half of the respondents believe that existing data systems for ordering essential health commodities are overly complex. These findings are consistent with the results of the previous study, which highlighted a shortage of health staff skilled in logistics supply chain management, leading to inefficient utilization of these data systems for departmental ordering [29]. Similarly, another study reported challenges in the availability of medicines, medical supplies, and reagents in many public health facilities in sub-Saharan countries, stemming from health workers’ limited ability to effectively use existing systems [28]. However, assessment of implementation of the health management information system at the district level in southern Malawi identified several factors contributing to the difficulty in using existing data systems for hospital staff allocation in low- and middle-income countries, including a shortage of skilled staff in Human Resource Information Systems for Health (HRIH) and other data sources [30] . These findings are supported by commendations from health care analysis that examined a strategy to improve priority setting in developing countries; that reported inadequate use of data systems and other data sources for health staff allocation in some health facilities due to a lack of training, resulting in resistance to data system adoption [31].

Furthermore, consistent with previous studies that highlighted the challenges faced by health staff in setting priorities and analyzing data during the planning process in developing countries—mainly due to limited capacity in using existing data systems for facility-level planning [32]. These findings align with the current study’s results, which indicate that a significant proportion of respondents found existing data systems insufficiently user-friendly for hospital planning and budget allocation, primarily due to inadequate staff training. In our study, challenges were also identified regarding the reliability of existing data systems for ordering essential health commodities, attributed to the limited capacity of health workers to use systems such as GoTHOMIS effectively. This is in line with a previous study, which found that modern health information technologies had a limited impact due to inadequate training in logistics supply chain management and insufficient supervisory support in health facilities, underscoring that underinvestment in training and supportive supervision weakens the effectiveness of digital systems for commodity ordering and management [33]. Additionally, the study findings show that a majority of respondents perceived existing data systems for allocating hospital staff at the departmental level as unreliable. These findings are supported by a study that discusses the chemical behaviour and stability of perovskite precursor inks, which argues that a key advantage of these materials is their straightforward solution processability, enabling low-cost and scalable fabrication [34].

Another study highlighted challenges faced by health staff in priority setting and using existing data systems during planning and budget allocation in low- and middle-income countries due to inadequate access to reliable information and weak institutional capacity [35]. These findings are consistent with the study’s results, which show that a considerable proportion of respondents viewed existing systems for analyzing and preparing hospital plans and budget allocation as unreliable, largely due to low staff capacity. The study findings also reveal that a significant proportion of respondents rated existing data systems as not useful for ordering essential health commodities. These findings align with, which reported that many primary health facilities still use paper-based integrated logistics management systems such as R&R forms, hindering effective data aggregation for informed decision-making on ordering medicines, medical supplies, and reagents [29].

A study a previous study indicated that many healthcare providers in primary health facilities did not use the available data sources for staff allocation due to limited knowledge and skills in integrated logistics management, leading to perceptions that existing data systems were ineffective for departmental staff allocation in health facilities [31]. These findings are supported by the study’s results, which show that a considerable proportion of respondents perceived existing data systems as not useful for staff allocation at departmental levels. Moreover, the study findings show that a substantial proportion of respondents rated existing data systems as not useful for preparing hospital plans and budget allocation at departmental and unit levels. These findings are consistent with another study that noted that limited capacity among health staff to analyses and prioritize using existing, often incomplete and inaccurate, modern technologies for planning and budget allocation contributed to low motivation and limited system use [35].

The study findings suggest that digital health factors—including the user-friendliness, reliability, and usefulness of existing data systems—play a crucial role in improving health service delivery by enabling better data use for planning, decision-making, and access to quality data. These results imply that strengthening the use of health data through effective adoption of digital health can substantially enhance service delivery in council hospitals. The findings also indicate that the compatibility of existing data systems has a positive effect on access to quality data. This is supported by evidence from annual public health reviews and analyses of health system performance, which show that the compatibility of health information technologies facilitates effective communication of technical information, including the visualization, sharing, dissemination, and interpretation of data in private health facilities [36].

Our findings are consistent with previous studies indicating that health information technology, particularly perceived ease of use, substantially influences access to quality data [9,12]. This suggests that the user-friendliness of health information systems positively affects access to quality data in council hospitals in Tanzania; when systems are intuitive and easy to navigate, users are more likely to adopt and routinely utilize them, thereby enhancing data availability and quality. Similarly, prior research has shown that nurses’ perceptions of the reliability of modern health information technologies shape their acceptance and use of these systems to access quality data through visualization and sharing with intended users in their facilities [8]. These observations align with our findings that the perceived reliability of existing data systems positively influences access to quality data in health facilities. When users have trust and confidence in the accuracy and stability of both the data and the systems that generate them, they are more likely to integrate these resources into routine decision-making, ultimately improving the quality of clinical and managerial decisions.

Furthermore, a previous study reported that the perceived usefulness of existing data has a positive effect on access to quality data, with perceived usefulness (path coefficient = 0.31, p<0.001) significantly strengthening behavioural intention to use digital systems [15]. Another study similarly demonstrated that perceived usefulness is a critical determinant in the adoption and effective use of information systems, particularly computerized systems, thereby influencing their use to access quality data [37]. These findings are in line with the present study, which shows that the perceived usefulness of existing data systems positively influences access to quality data, thereby supporting hypothesis H4 (β = 0.369, p<0.001). Collectively, these results suggest that when users perceive a system as useful for accomplishing their tasks, they are more likely to engage with it, which in turn enhances access to quality data from existing data systems. However, a study on the factors affecting the adoption of healthcare found that users tend to use information systems more intensively when these systems provide timely, complete, and accurate outputs that meet their information needs and support goal attainment [38]. These findings imply that adequate access to quality data through existing data systems, coupled with appropriate use of health data and supported by modern information technologies, can improve health service delivery in low- and middle-income countries, including Tanzania. They are also consistent with evidence indicating that access to quality data positively influences data use for planning and decision-making, thereby supporting hypothesis H9 (β = 0.729, p<0.001).

The study findings demonstrate that data use for planning and decision-making has a positive and significant effect on improving health service delivery. These results suggest that hospitals with effective health management information systems are better able to empower staff to use health data, thereby enhancing the quality of care in their workplaces. The findings are consistent with a previous study [39] and with the UTAUT model [16], which posits that Performance Expectancy, Effort Expectancy, and Social Influence positively affect behavioural intention to use information technology, while facilitating conditions positively affect actual technology use. Similarly, a national study on the hospital computing and the costs and quality of care reported that hospital staff were more likely to use existing data systems when management emphasized their importance, which in turn promoted effective health data use and contributed to improved healthcare within their facilities [40].

## Limitations of the study

A key limitation of this study is that it focused exclusively on members of Quality Improvement teams and did not include other relevant groups, such as Health Management Teams and Workplace Improvement Teams operating in council hospitals. The exclusion of these groups may have restricted the breadth of perspectives on how health information technology influences health data use within the studied facilities. Furthermore, the study was limited to council hospitals and did not encompass dispensaries, health centres, regional referral hospitals, national hospitals, or specialized hospitals. As a result, it remains unclear whether the patterns of health data use for decision-making observed in council hospitals are similar to, or differ from, those in other levels of the health system, thereby leaving an important gap and limiting the generalizability of the findings.

## Conclusion

The study concludes that, in their current form, digital health initiatives have had a limited overall influence on the user-friendliness, reliability, and perceived usefulness of existing data systems for accessing quality data and supporting data use for planning and decision-making in council hospitals. The coexistence of multiple, poorly integrated systems with limited interoperability has hindered data exchange, generated inconsistencies, and increased the burden of redundant data management, thereby constraining effective health data use. At the same time, the findings demonstrate that specific digital health factors—particularly compatibility, user-friendliness, reliability, and usefulness—have a positive and statistically significant effect on improving health service delivery through enhanced access to quality data and more systematic data use for planning and decision-making. This suggests that increasing acceptance and effective use of digital health can foster greater and more meaningful use of data, ultimately contributing to improved health service delivery in council hospitals. Accordingly, the study recommends that government authorities strengthen existing data systems, promote their integration into a unified data warehouse to facilitate seamless data exchange and use, and provide comprehensive, continuous training on digital health for all relevant cadres. Such measures are essential to maximize the benefits of digital health investments and to institutionalize a culture of data-informed decision-making within the health system.

## Authors’ Contributions

Author contributions were as follows: Constantine Matimo and Henry Mollel contributed to the conceptualization of the study. Formal analysis was conducted by Constantine Matimo and Godfrey Kacholi. Constantine Matimo was responsible for the investigation. The methodology was developed by Constantine Matimo and Henry Mollel. Supervision was provided by Henry Mollel. Validation was undertaken by Constantine Matimo. The original draft of the manuscript was written by Constantine Matimo, Godfrey Kacholi, and Henry Mollel, and all authors contributed to writing, review, and editing. All authors have read and approved the final version of the manuscript.

## Data Availability

No datasets were generated or analyzed during the current study. All relevant data from this study will be made available upon study completion.

## Acknowledgements

The authors express their sincere gratitude to the President’s Office – Regional Administration and Local Government, as well as the Regional Administrative and Local Authorities, for granting permission to conduct this study in the selected council hospitals. We also acknowledge the valuable cooperation, contributions, and logistical support provided by the district medical officers and medical officers in charge of the surveyed council hospitals. The authors are especially grateful to the members of the QI teams in the participating council hospitals for generously providing the information that made this study possible.

